# The longitudinal relation between executive functioning and multilayer network topology in glioma patients

**DOI:** 10.1101/2022.07.22.22277928

**Authors:** Marike R. van Lingen, Lucas C. Breedt, Jeroen J.G. Geurts, Arjan Hillebrand, Martin Klein, Mathilde C.M. Kouwenhoven, Shanna D. Kulik, Jaap C. Reijneveld, Cornelis J. Stam, Philip C. De Witt Hamer, Mona L.M. Zimmermann, Fernando A.N. Santos, Linda Douw

## Abstract

Many patients with glioma, primary brain tumors, suffer from poorly understood executive functioning deficits before and/or after tumor resection. We aimed to test whether frontoparietal network centrality of multilayer networks, which allow for integration across multiple frequencies, relates to and predicts executive functioning in glioma patients before and after tumor resection. Patients with glioma (n = 37) underwent neuropsychological tests assessing word fluency, inhibition, and set shifting, and resting-state magnetoencephalography before tumor resection (T1) and one year after resection (T2). We constructed binary multilayer networks comprising six layers, with each layer representing frequency-specific functional connectivity (phase lag index) between source-localized time series of 78 cortical regions. Average frontoparietal network multilayer eigenvector centrality, a measure for network integration, was calculated at both time points. Regression analyses were used to investigate its associations with executive functioning.

At T1, lower multilayer integration (*p =* 0.017) and having epilepsy (*p =* 0.006) associated with poorer set shifting (adj. *R*^*2*^ = 0.269). Decreasing multilayer integration (*p =* 0.022) and not undergoing chemotherapy at T2 (*p =* 0.004) related to deteriorating set shifting (adj. *R*^*2*^ = 0.283). No significant associations were found for word fluency or inhibition, nor did T1 multilayer integration predict changes in executive functioning. As expected, our results establish multilayer integration of the frontoparietal network as a cross-sectional and longitudinal correlate of executive functioning in glioma patients. However, multilayer integration did not significantly predict postoperative changes in executive functioning, limiting its direct clinical relevance.

## Introduction

Gliomas, originating from glial cells, are the most common primary brain tumors and have a fatal disease course. Despite their local appearance on MRI, patients experience varied cognitive complaints that cannot be attributed to location alone (De Baene et al., 2019; van Kessel et al., 2017). Executive functioning (EF), is often affected at diagnosis, before treatment starts (Noll et al., 2020; Tanzilli et al., 2022; van Kessel et al., 2017), and may contribute to lower quality of life (Weyer-Jamora et al., 2021). Subsequently, there is large individual variability in cognitive trajectories across the disease course. After tumor resection, improving, stable, as well as deteriorating EF has been observed (Lemaitre et al., 2021; Ng et al., 2019; Noll et al., 2015; Satoer et al., 2012; Sinha et al., 2020; Talacchi et al., 2011; Wu et al., 2011). Many patients also receive chemo- and/or radiotherapy, depending on tumor subtype and residual tumor, which also impacts EF both positively and negatively according to the literature (Hilverda et al., 2010; Koutsarnakis et al., 2021; Tabor et al., 2021; Talacchi et al., 2011; Tanzilli et al., 2022). Other relevant correlates of poorer EF are higher age, lower Karnofsky performance status (KPS (Karnofsky et al., 1948)), frontal tumor location (Fang et al., 2014), and use of antiepileptic drugs (Klein et al., 2004). However, it remains impossible to predict individual cognitive trajectories with reasonable accuracy, leading to uncertainty about future cognitive performance.

The currently known neural correlates of EF mainly comprise connectivity and network-based variables, reflecting the distributed brain networks involved. Functional brain connectivity refers to the statistical interdependencies between brain activity of different network nodes or brain regions (Aertsen et al., 1989; Friston, 1994). Network theory can then further assess local and global properties of this brain network (Bassett & Sporns, 2017; Sporns et al., 2005; Stam & Reijneveld, 2007; van den Heuvel & Hulshoff Pol, 2010). Cognition in general, and EF particularly, has been shown to depend on long-distance integration across distributed brain regions, which can be operationalized through the centrality of regions within cognitively relevant networks (Baum et al., 2017; Deco et al., 2021; Medaglia et al., 2015; Sauseng et al., 2005).

In glioma, functional MRI-based connectivity of the frontoparietal network (FPN) is relevant for EF (Kocher et al., 2020; Landers et al., 2021; Maesawa et al., 2015; Noll et al., 2015, 2021; Tordjman et al., 2021). Using magnetoencephalography (MEG), frequency-specific connectivity has also been linked to EF in these patients: poorer EF at diagnosis relates to lower alpha band (8-13Hz) functional connectivity (Derks et al., 2019), and particularly lower integrative connectivity in the theta (4-8Hz), alpha, and beta (13-30Hz) bands associate with poorer EF cross-sectionally (Bosma et al., 2009) and longitudinally (Carbo et al., 2017; van Dellen et al., 2012a).

Previous studies constructed separate functional networks for each frequency band, which may relate to particular cognitive aspects, e.g. alpha band oscillations to attention (Klimesch, 2012), gamma band to memory (Kucewicz et al., 2014), and beta band to sensorimotor functions (Kilavik et al., 2013). In EF, all these cognitive aspects are combined, rendering it a relevant domain for frequency-integrated investigations. The multilayer network approach can be used to synergize frequencies, by defining a network within each frequency (layer), and then coupling the different layers (De Domenico et al., 2013). Nodes representing the same brain regions are present in each layer (Brookes et al., 2016; De Domenico et al., 2016; Guillon et al., 2017; Tewarie et al., 2016). Indeed, lower multilayer centrality of the FPN (including MEG and MRI layers) related to poorer EF in healthy subjects (Breedt et al., 2021). Associations between cognition and MEG multilayer centrality were also found in Alzheimer’s disease (Yu et al., 2017).

In this study, we tested the hypotheses that (1) lower multilayer FPN integration correlates with poorer EF at diagnosis, and (2) changes in EF and multilayer centrality coincide. Finally, we hypothesized (3) lower multilayer centrality at diagnosis to predict deteriorating EF after tumor resection.

## Methods

### Patients

Consecutive new patients between 2011-2021 at Amsterdam UMC with suspected diffuse glioma were eligible for participation in an ongoing prospective study on brain networks. Exclusion criteria were (1) age <18 years, (2) psychiatric disease, (3) comorbidities of the central nervous system, (4) insufficient mastery of the Dutch language, and (5) inability to communicate adequately. After resection, molecular characteristics were assessed as part of clinical routine, including prognostically favorable isocitrate dehydrogenase (IDH) mutations and 1p/19q codeletions (Louis et al., 2021). This led to three subgroups: IDH-wildtype glioma (glioblastoma), IDH-mutant, non-codeleted glioma, and IDH-mutant, 1p/19q-codeleted glioma.

Patients underwent neuropsychological assessments (NPA (Derks et al., 2019)) and MEG at two time points: preoperatively at diagnosis (T1), and approximately one year after tumor resection (T2). Functional and diffusion MRI and patient-reported outcomes were acquired, but not included in the current work. At T2, MEG and NPA took place between 8-20 months after tumor resection, with a maximum of 3 months between them.

Part of this data has been reported on previously (Belgers et al., 2020; Carbo et al., 2017; Derks et al., 2018, 2019, 2021; Douw et al., 2008; van Dellen et al., 2012a, 2012b). The current analysis was preregistered before selecting eligible patients and performing analyses (https://osf.io/83tbq).

The VUmc Medical Ethical Committee approved this study, which was conducted following the principles of the Declaration of Helsinki. All participants provided written informed consent prior to participation.

### Neuropsychological assessment

Three EF tests were performed: the Categoric Word Fluency test (Mulder et al., 2006) for planning and strategy, the Concept Shifting Test (van der Elst et al., 2006) for attention, working memory and set shifting, and the Stroop Color-Word Test (Hammes, 1978) for attention and inhibition (see Supplementary Materials). In short, each test yielded a single final score, which was adjusted for age, sex, and educational level and converted to a Z-score using validated normative data. The commonly used cut-off value of Z < -1.5 was used to indicate cognitive impairment (Sinha et al., 2020).

### Magnetoencephalography

MEG was recorded for 5min in supine position during eyes closed no-task resting-state in a magnetically shielded room (VacuumSchmelze GmBh, Hanau, Germany), using a 306-channel (102 magnetometers and 204 gradiometers) whole-head MEG system (Elekta Neuromag Oy, Helsinki, Finland) and a sampling frequency of 1250Hz. Anti-aliasing and high-pass filters of 410 and 0.1Hz, respectively, were applied online.

In short, preprocessing involved visual inspection, removal of noisy channels, and noise removal in the remaining signals (see Supplementary Materials for details). Anatomical MRI was used for co-registration with the digitized scalp surface, and the Automated Anatomical Labeling atlas (Tzourio-Mazoyer et al., 2002) was used to parcellate the cortical ribbon into 78 brain regions.

Broadband time series of neuronal activity were then reconstructed for each region’s centroid (Hillebrand et al., 2016) using a scalar beamformer approach (Hillebrand et al., 2012). For each patient, we included the first 60 epochs of 4096 samples (3.28s; total >3min). Fast Fourier transforms filtered the time series into six classical frequency bands: delta (0.5-4Hz), theta (4-8Hz), lower alpha (8-10Hz), upper alpha (10-13Hz), beta (13-30Hz), and gamma (30-48Hz). Finally, we computed and averaged the phase lag index (PLI (Stam et al., 2007)) between the frequency-filtered time series of region pairs using custom-made scripts in MATLAB (R2020b, MathWorks, Natick, MA, USA; see https://github.com/multinetlab-amsterdam for associated code and data), yielding a single weighted network per frequency band per patient.

### Multilayer network analysis

We used the multiplex network (Bianconi, 2018), which contains interlayer links only between the same nodes or brain regions across layers. Since differences in weight distribution across network layers impact multilayer network topology (Mandke et al., 2018), we first binarized each layer separately through Kruskal’s algorithm to construct minimum spanning trees (MSTs (Stam et al., 2014; Tewarie et al., 2015)). The MST recapitulates the network’s backbone, by incorporating each node and optimizing the overall weight of the network with n-1 connections (in our case 77 connections) and no cycles. The MST has been amply applied in MEG, with a recent meta-analysis revealing consistent transdiagnostic MST alterations (Blomsma et al., 2022). We set all interlayer link weights to 1 (Figure 1).

**Figure 1.**
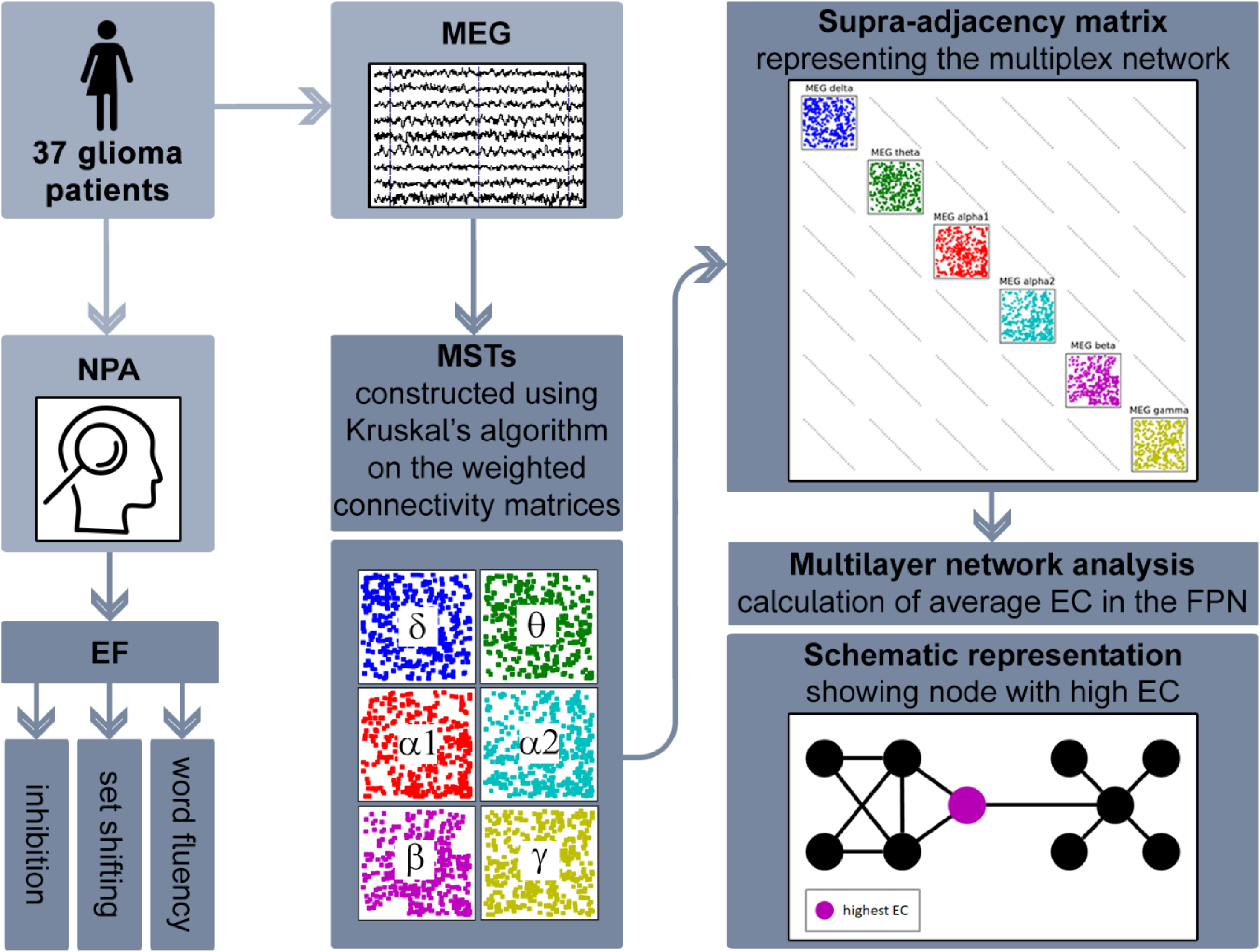
Schematic overview of the MEG multilayer analysis pipeline. For every participant, magnetoencephalography (MEG) data was preprocessed and projected to the brain; the brain was parcellated according to the Automated Anatomical Labeling (AAL) atlas; the Phase Lag Index (PLI) was used to compute weighted connectivity matrices; minimum spanning trees (MST) of the weighted matrices were constructed using Kruskal’s algorithm; and finally an LxN by LxN supra-adjacency matrix representing a multilayer network of the six MEG frequency bands as layers (all with N = 78 nodes for each AAL region and M = N – 1 = 77 binary intralayer links) was constructed, where diagonal blocks contain the intralayer connections for each frequency band and the off-diagonal blocks the interlayer connections; like the intralayer connections, we set all interlayer link weights to 1, obtaining binary multilayer networks; now, multilayer eigenvector centrality (EC) of the frontoparietal network (FPN) was calculated and averaged for each patient and timepoint. NPA = neuropsychological assessment, EF = executive functioning.

We then calculated nodal multilayer eigenvector centrality (EC) as a measure of integration according to De Domenico et al. (2016) in Python (version 3.6, Python Software Foundation). EC not only takes the number of connections a node itself has into account, but also considers the centrality of neighboring nodes (Lohmann et al., 2010). EC relates to cognition in MEG literature (Hardmeier et al., 2012) and to EF in particular when using the multilayer approach (Breedt et al., 2021). Finally, nodal EC values of FPN nodes (according to (van Dellen et al., 2012a); Supplementary Table 2) were averaged, resulting in one multilayer FPN EC value per patient per time point.

### Statistical analysis

Statistical analyses were performed using IBM SPSS (version 26.0, IBM Corp, Armonk, NY, USA). Paired t-tests (or Wilcoxon signed-rank tests in case of non-normality) were used to assess cognitive and network changes.

Backward linear regressions tested whether multilayer EC associated with EF at T1. For each EF aspect, a separate regression was performed. Based on literature (Derks et al., 2019; van Kessel et al., 2017; Wefel et al., 2016), glioma subtype and tumor location were selected as covariates. Potential additional covariates were selected through significant (*p* < 0.05) associations with the dependent variable (through correlation coefficients for age, time between resection and NPA/MEG, tumor volume, percent tumor overlap with FPN; through t-tests or Mann-Whitney U tests for tumor lateralization, presence of epilepsy, education, sex, handedness, active chemotherapy treatment during or <4 weeks before T2; through (Kruskal-Wallis) ANOVA for tumor grade, treatment type). Patients with tumor progression before T2 were only included for this cross-sectional T1 analysis.

To test whether multilayer EC changes (T2-T1) related to cognitive changes (T2-T1), backward linear regressions were performed. Based on literature (Hilverda et al., 2010; Koutsarnakis et al., 2021; Tabor et al., 2021; Talacchi et al., 2011), type of anti-tumor treatment (radiotherapy, chemotherapy, chemoradiation, or no treatment) was included as coviarate. We initially planned on performing longitudinal analyses separately in patients with progression before T2, but this sample proved too small.

Backward linear regressions were used to test whether T1 multilayer EC predicted changes in EF. Again, anti-tumor treatment was included as covariate, and active chemotherapy was explored as a potential additional covariate.

The level of significance was set at *p* < 0.05, but significance after Bonferroni correction was also reported (*p*-value multiplied by three, for each EF aspect).

## Results

### Patient characteristics

In total, 37 participants (mean age 41.7 years ± SD 12.3) completed NPA and MEG at T1 and T2 (Table 1). For two patients, the T1 set shifting score could not be calculated due to missing motor scores; three patients had incomplete T2 inhibition scores; one patient had a missing T2 word fluency score. In 4 patients, the interval between T2 MEG and NPA was >3 months (range 4-7), but they were included as they had stable disease for at least 17 months after surgery.

**Table 1.**
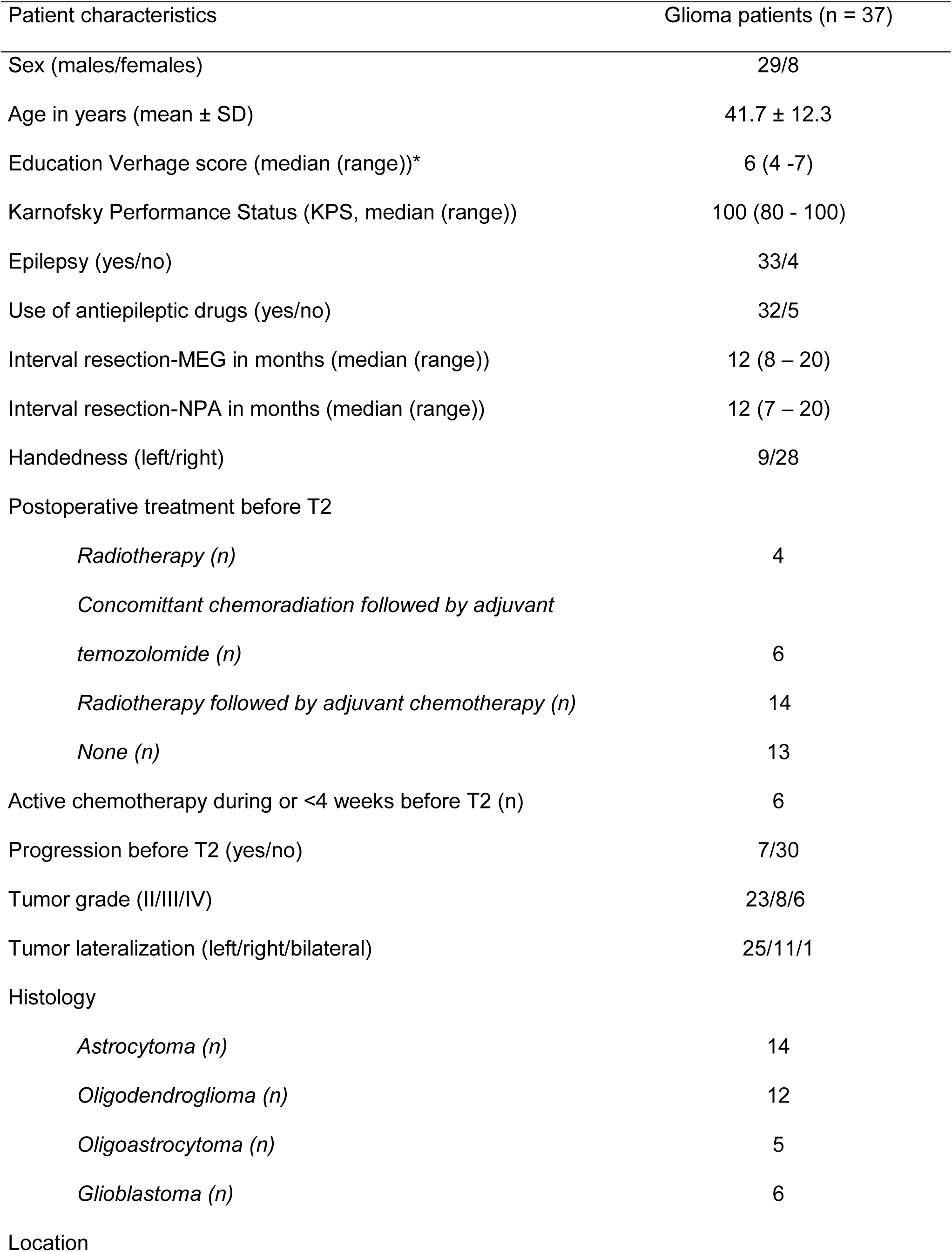
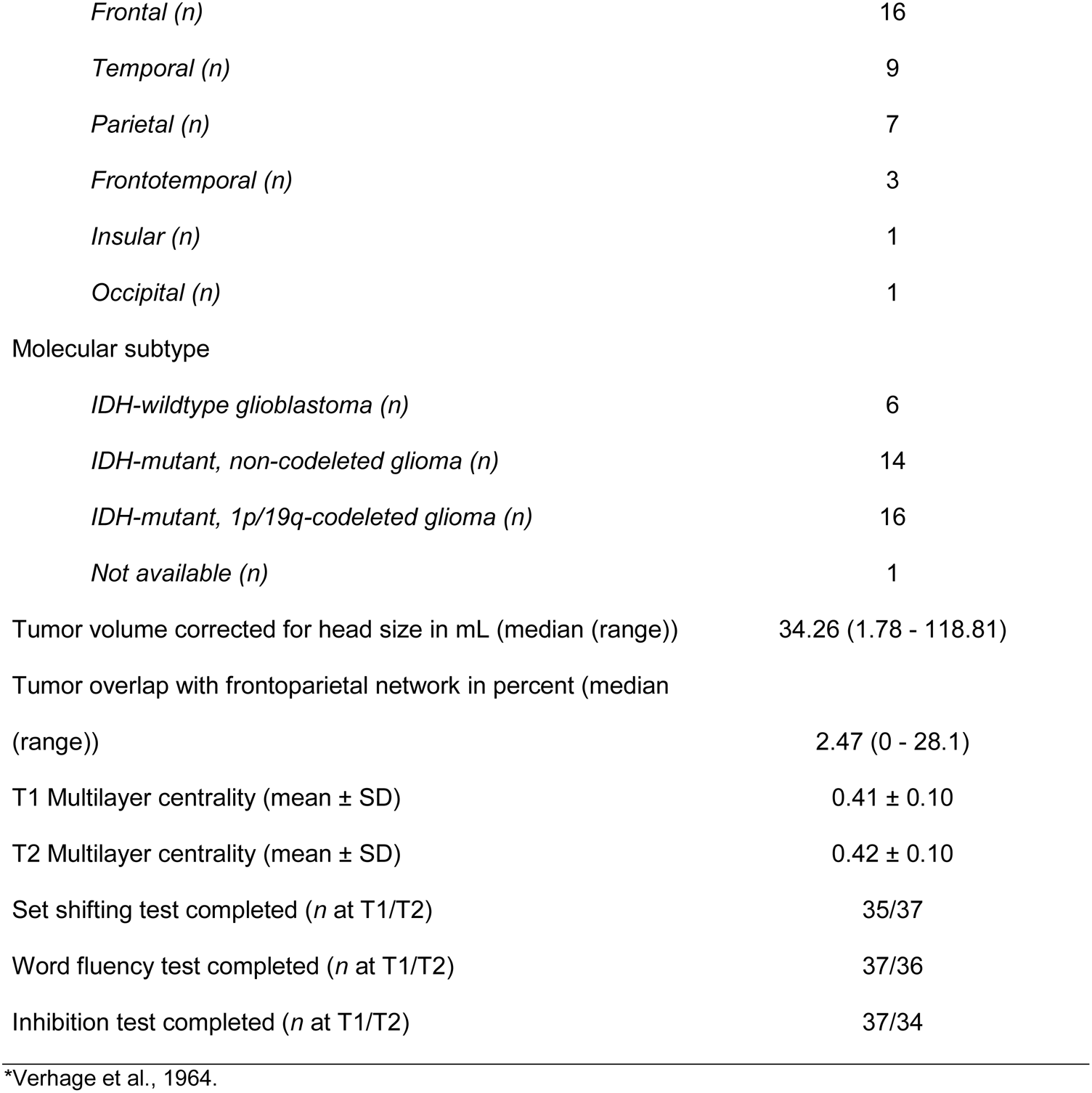
Patient characteristics

| Patient characteristics | Glioma patients (n = 37) |
| --- | --- |
| Sex (males/females) | 29/8 |
| Age in years (mean ± SD) | 41.7 ± 12.3 |
| Education Verhage score (median (range))* | 6 (4 -7) |
| Karnofsky Performance Status (KPS, median (range)) | 100 (80 - 100) |
| Epilepsy (yes/no) | 33/4 |
| Use of antiepileptic drugs (yes/no) | 32/5 |
| Interval resection-MEG in months (median (range)) | 12 (8 – 20) |
| Interval resection-NPA in months (median (range)) | 12 (7 – 20) |
| Handedness (left/right) | 9/28 |
| Postoperative treatment before T2 |  |
| *Radiotherapy (n)* | 4 |
| *Concomittant chemoradiation followed by adjuvant temozolomide (n)* | 6 |
| *Radiotherapy followed by adjuvant chemotherapy (n)* | 14 |
| *None (n)* | 13 |
| Active chemotherapy during or <4 weeks before T2 (n) | 6 |
| Progression before T2 (yes/no) | 7/30 |
| Tumor grade (II/III/IV) | 23/8/6 |
| Tumor lateralization (left/right/bilateral) | 25/11/1 |
| Histology |  |
| *Astrocytoma (n)* | 14 |
| *Oligodendroglioma (n)* | 12 |
| *Oligoastrocytoma (n)* | 5 |
| *Glioblastoma (n)* | 6 |
| Location |  |
| *Frontal (n)* | 16 |
| *Temporal (n)* | 9 |
| *Parietal (n)* | 7 |
| *Frontotemporal (n)* | 3 |
| *Insular (n)* | 1 |
| *Occipital (n)* | 1 |
| Molecular subtype |  |
| *IDH-wildtype glioblastoma (n)* | 6 |
| *IDH-mutant, non-codeleted glioma (n)* | 14 |
| *IDH-mutant, 1p/19q-codeleted glioma (n)* | 16 |
| *Not available (n)* | 1 |
| Tumor volume corrected for head size in mL (median (range)) | 34.26 (1.78 - 118.81) |
| Tumor overlap with frontoparietal network in percent (median (range)) | 2.47 (0 - 28.1) |
| T1 Multilayer centrality (mean ± SD) | 0.41 ± 0.10 |
| T2 Multilayer centrality (mean ± SD) | 0.42 ± 0.10 |
| Set shifting test completed (*n* at T1/T2) | 35/37 |
| Word fluency test completed (*n* at T1/T2) | 37/36 |
| Inhibition test completed (*n* at T1/T2) | 37/34 |

\*Verhage et al., 1964.

At T1, 8 patients (22%) showed impaired set shifting (Figure 2A); 2 patients (5%) had impaired word fluency; no patients had impaired inhibition. Longitudinally, 4 patients (11%) became unimpaired and 3 patients (8%) became impaired in set shifting; 4 patients (11%) declined to impaired word fluency; 1 patient (3%) became impaired in inhibition.

**Figure 2.**
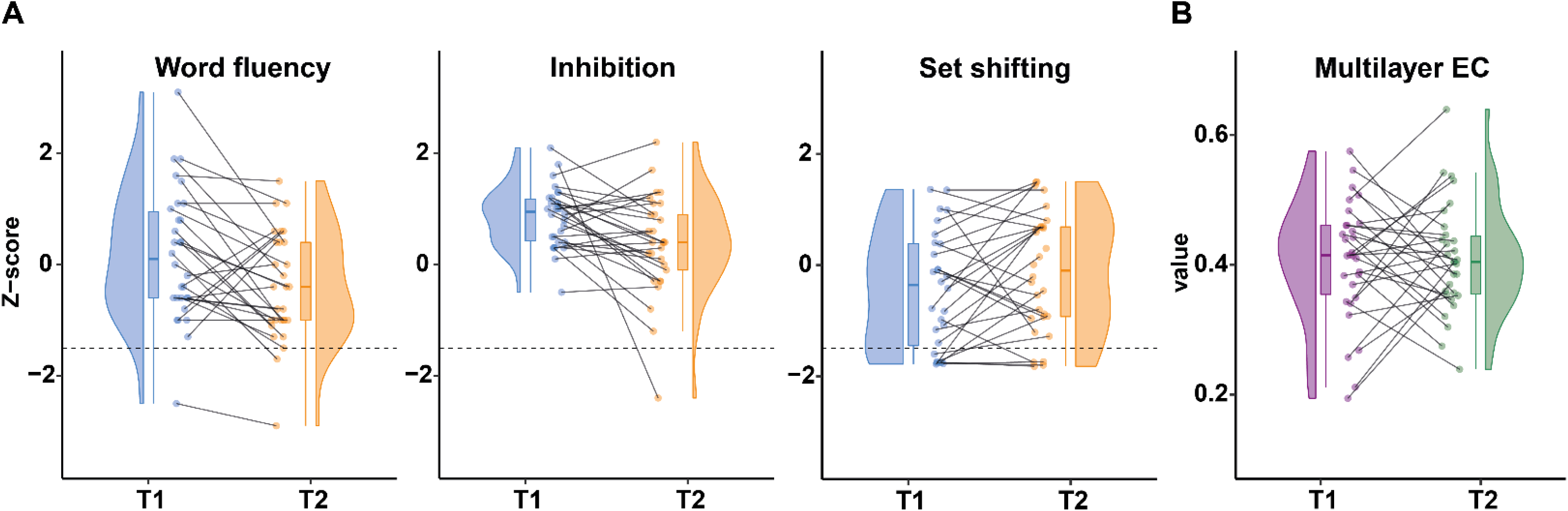
Executive functioning and multilayer integration at both time points. Each panel shows a paired raincloud plot, in which individual data points of each patient at both time points are displayed through the combination of a scatterplot (the ‘rain’), a spaghetti plot, a box plot, and a probability density plot (the ‘cloud’). Panel A shows patients’ Z-scores of the three executive functioning tests. Scores below the dashed line at -1.5 indicate clinically relevant cognitive deficits. Only word fluency changed significantly at the group-level (corrected *p* = 0.006). Panel B shows patients’ multilayer eigenvector centrality (EC) of the frontoparietal network, which did not change significantly at the group-level.

At the group-level, word fluency declined significantly (uncorrected *p* = 0.002, corrected *p* = 0.006, Figure 2A). Inhibition declined non-significantly (uncorrected *p* = 0.057). Set shifting remained unchanged (uncorrected *p =* 0.441).

At the group-level, multilayer EC did not change (uncorrected *p =* 0.755, Table 1 and Figure 2B, Supplementary Figure 2 visually represents molecular subtypes separately).

**Table 2.**
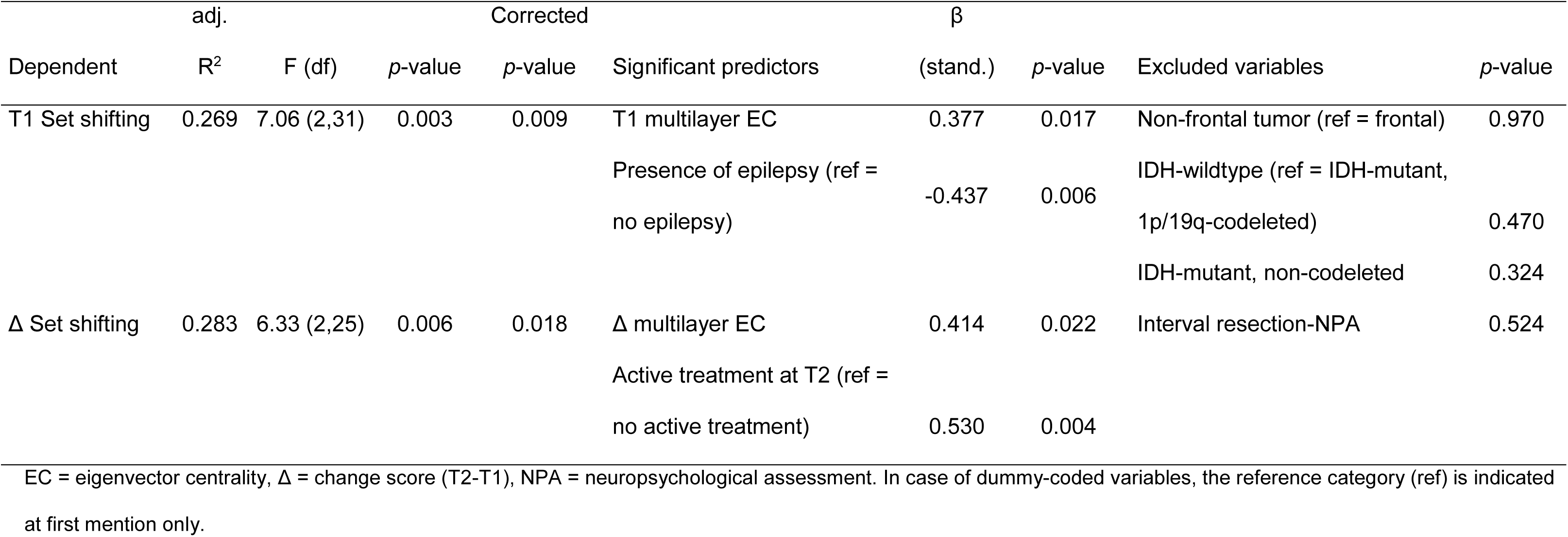
Significant regression results concerning multilayer integration

### Baseline brain-cognition correlations

For set shifting, having epilepsy or not was included as an additional covariate at T1, whereas no additional covariates were selected for word fluency and inhibition. Lower multilayer EC and having epilepsy significantly related to poorer set shifting (*p* = 0.017 and *p* = 0.006 respectively, model corrected *p* = 0.009; Table 2, Figure 3A). Multilayer EC was not included in the final models for word fluency and inhibition, but patients with IDH-mutant, 1p/19q-codeleted glioma had better word fluency than IDH-wildtype glioblastoma patients (Supplementary Table 2).

**Figure 3.**
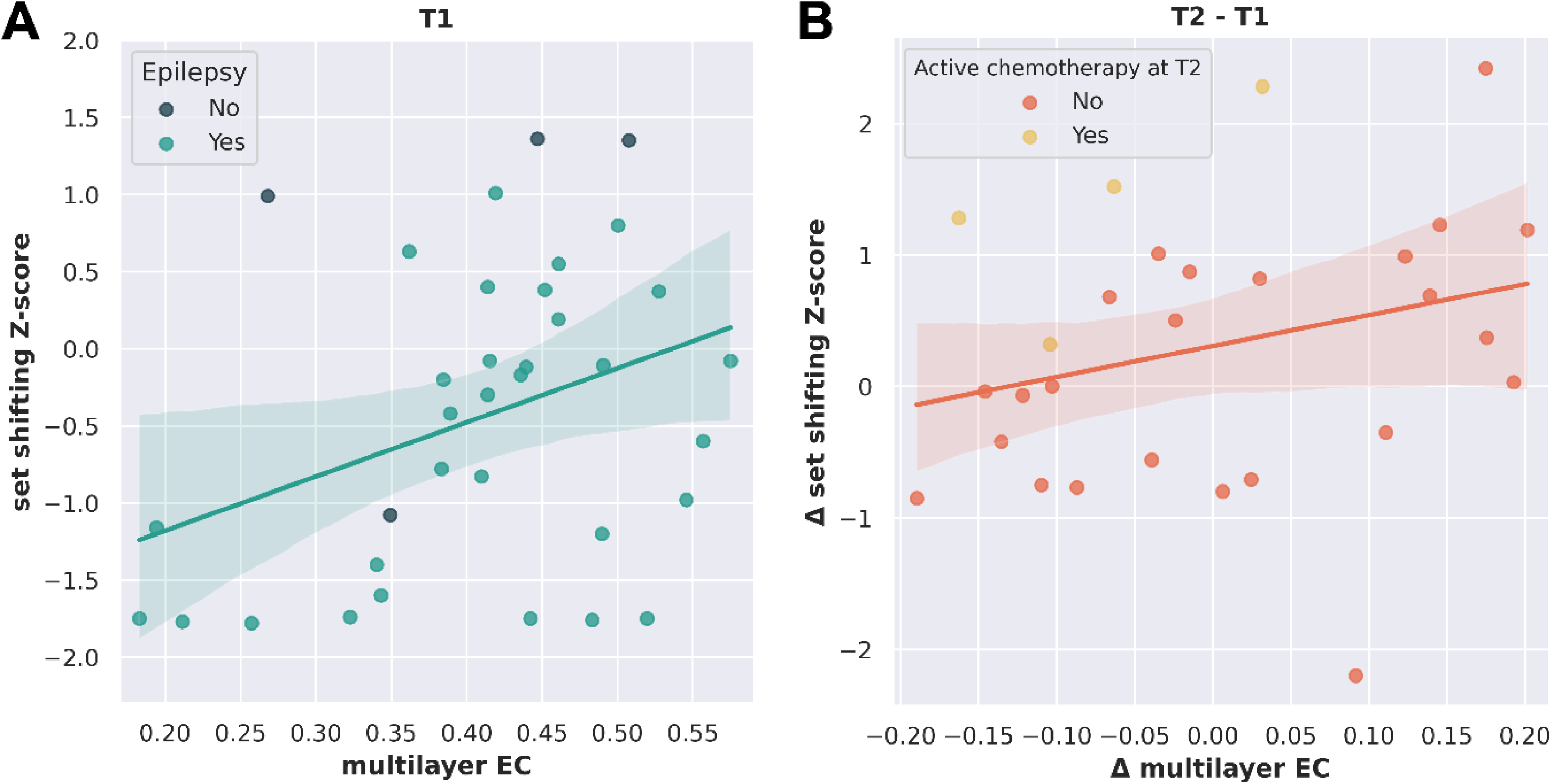
Significant associations between multilayer integration and set shifting. (A) Displays the cross-sectional association between multilayer eigenvector centrality (EC) of the frontoparietal network and set shifting performance (n = 35, model corrected *p* = 0.009), with color codes indicating the included covariate (presence of epilepsy). (B) Shows the longitudinal association between change in multilayer EC (T2-T1) and change in set shifting (T2-T1; n = 28, model corrected *p* = 0.018), with color codes indicating the included covariate (active chemotherapy at T2). Shaded area: 95% confidence interval.

### Longitudinal brain-cognition correlations

Longitudinal regression analyses were performed in patients without tumor progression before T2 (n = 30). For set shifting, type of anti-tumor treatment, active chemotherapy, resection-NPA interval, and resection-MEG interval correlated with change scores. Due to multicollinearity, only active treatment and resection-NPA interval were included. Decreasing multilayer EC and no active chemotherapy were associated with declining set shifting (*p =* 0.022 and *p =* 0.004, respectively, model corrected *p =* 0.018; Table 2, Figure 3B).

Multilayer EC changes did not relate to changes in inhibition and word fluency (Supplementary Table 1). Patients with IDH-mutant, 1p/19q-codeleted glioma showed greater word fluency decline than patients with IDH-mutant, non-codeleted glioma (*p =* 0.001, Supplementary Figure 1) and IDH-wildtype glioblastoma (*p =* 0.005, model corrected *p =* 0.003).

### Baseline predictors of cognitive change

T1 multilayer EC did not predict cognitive change (Supplementary Table 1). Instead, poorer baseline inhibition and word fluency were associated with improving performance (*p =* 0.014 and *p* < 0.001, respectively, model corrected *p* = 0.066 and *p* < 0.001, respectively). For set shifting, poorer baseline performance and active chemotherapy were related to improving scores (model corrected *p* = 0.049), but individual predictors did not reach significance (*p* = 0.069 and *p* = 0.091, respectively).

## Discussion

As expected, a relevant proportion of glioma patients showed executive dysfunction in this study, both at diagnosis and after tumor resection. While there were group-level decreases in word fluency only, we found variable individual trajectories for inhibition and set shifting. Partly confirming our hypotheses, lower and decreasing multilayer FPN integration (eigenvector centrality) related to poorer and deteriorating set shifting, but not word fluency and inhibition. Contrary to our hypothesis, baseline multilayer centrality did not predict postoperative changes in executive functioning.

Our findings on the association between multilayer integration and set shifting are in line with previous findings in healthy controls (Breedt et al., 2021), even though the study by Breedt used a composite EF score. Central nodes like those within the FPN are thought to facilitate global communication between segregated communities, presumably enabling EF (Bertolero et al., 2017; Sporns, 2013). Our multilayer analysis indeed picks up on relevant individual variation in EF in these patients, explaining ∼27% of the cognitive variance in set shifting at diagnosis. Furthermore, our findings synthesize previous frequency-specific findings in relation to cognitive functioning in these patients (Bosma et al., 2009; Carbo et al., 2017; van Dellen et al., 2012a). These findings may particularly inform future studies on the development of targeted cognitive treatment. For example, non-invasive stimulation of the FPN improves cognitive functioning in healthy adults (Luber & Lisanby, 2014), with a recent preliminary study demonstrating positive neurological results in glioma patients (Poologaindran et al., 2022). Baseline functional network properties may help determine the efficacy of stimulation and its subsequent effect on cognition (Douw et al., 2020; Fitzsimmons et al., 2020; Nicolo et al., 2015; Williams et al., 2021); multilayer FPN integration may thus be explored as a target in individual patients.

Correlations were significant only for set shifting, potentially reflecting particular sensitivity of multilayer integration towards this aspect of EF. Of note, performance on the three tests showed very different distributions, and the large variability in set shifting performance over time may have led to greater statistical power to find correlations with multilayer integration. Of course, the lack of findings for inhibition and word fluency could also be due to the small and heterogeneous sample. In particular, although we statistically adjusted for molecular subtype, scores on especially word fluency diverged between these subtypes. Moreover, poorer test scores at T1 predicted larger improvements in both word fluency and inhibition, potentially reflecting regression to the mean. In terms of covariates, having epilepsy was related to poorer set shifting at T1, congruent with earlier work (Klein et al., 2003). The finding that patients undergoing active chemotherapy at T2 showed improving set shifting remains difficult to interpret, particularly since none of these patients had shown tumor progression yet.

Contrary to our hypothesis, we did not find predictive significance of baseline multilayer integration towards EF change. Our current results suggest that although relevant as a cross-sectional EF correlate, multilayer integration as operationalized here may not predict EF trajectories in a heterogeneous sample of glioma patients.

There are several limitations that should be taken into account when interpreting these results. Firstly, although large for a rare disease, our sample size is still relatively small. Furthermore, we included a heterogeneous cohort, while molecular subtype particularly may pertain to cognitive functioning (Derks et al., 2019; Wefel et al., 2016). Future studies should include more patients of each subtype to enable more specific assessment of brain-cognition correlations, instead of merely adjusting analyses for subtype. Thirdly, participation bias and therefore generalizability should always be considered in observational studies, as supported by our patients’ high performance status. Fourthly, the multilayer network approach is new; other choices in terms of defining intra- and interlayer connectivity (Boccaletti et al., 2014) could be explored now that we established that multilayer integration relates to EF in these patients. Finally, we used a relatively coarse and non-connectivity-based atlas to allow for comparison with earlier work (Carbo et al., 2017; Derks et al., 2019; van Dellen et al., 2012a, 2012b), but replication with a connectivity-based atlas may yield additional insightful results.

## Conclusion

We describe multilayer frequency-band integration of the frontoparietal network as a new correlate of executive functioning in glioma patients. Lower and decreasing integration related to poorer and declining set shifting. Unfortunately, baseline multilayer centrality did not have predictive value for declining EF over time. Still, our findings may inform future studies on the brain mechanisms underlying cognitive decline in these patients, as well as spark exploration of more targeted treatment of EF deficits, for instance through non-invasive brain stimulation based on a multilayer target.

## Data Availability

This analysis on existing data has been preregistered (https://osf.io/83tbq). Derivative data needed to replicate these results can be found at https://github.com/multinetlab-amsterdam.
Scripts that were used to construct the supra-adjacency matrices in MATLAB and multilayer eigenvector centrality in the frontoparietal network in Python can be found here:
https://github.com/multinetlab-amsterdam

https://github.com/multinetlab-amsterdam

## Ethics approval

Approval for this study was obtained from the VUmc Medical Ethical Committee.

## Consent to participate

Informed consent was obtained from all individual participants included in the study.

## Consent for publication

(include appropriate statements)

## Availability of data and material

This analysis on existing data has been preregistered (https://osf.io/83tbq). Derivative data needed to replicate these results can be found at https://github.com/multinetlab-amsterdam.

## Code availability

Scripts that were used to construct the supra-adjacency matrices in MATLAB and multilayer eigenvector centrality in the frontoparietal network in Python can be found here: https://github.com/multinetlab-amsterdam

## Author contributions

included conception and study design (MRvL, LD), data collection or acquisition (MRvL, SDK), preprocessing of MEG data (LCB, SDK, MLMZ), statistical analysis (MRvL, LD, LCB, FANS), interpretation of results (MRvL, LD, LCB), drafting the manuscript work or revising it critically for important intellectual content (all authors) and approval of final version to be published and agreement to be accountable for the integrity and accuracy of all aspects of the work (all authors).

