## Supplementary Materials for "The longitudinal relation between executive functioning and multilayer network topology in glioma patients"

### Methods

#### *Neuropsychological assessment*

The goal of the Word Fluency Test (WFT (Mulder et al., 2006)) is for the participant to list, within 60 seconds, as many words as they can in a particular category. In the present study, we used the category 'animals'.

The Concept Shifting Test (CST (Van der Elst et al., 2006)) comprises 16 small circles all of which contain a digit (part A), a letter (part B), or a letter or a digit (part C). The small circles are grouped into a larger circle. The goal of the test is for the participant to cross out the circles as fast as possible without making errors, in ascending (part A), alphabetical (part B), or alternating order (digit-letter; part C). To correct for motor speed, the participant additionally performs a dummy condition thrice, with empty circles.

The Stroop Color-Word Test (SCWT (Hammes, 1978)) consists of three cards which have to be read aloud by the participant. The first card contains the names of four colors (red, green, yellow, and blue) printed in black ink. The second card contains rectangles printed in these same colors. The third card contains the names of the four colors from the first card, but printed in an inconsistent color ink (e.g., the word 'blue' is printed in red ink) and the participant is told to ignore the word and list the color of the ink. All cards have to be read as fast as possible, without making mistakes.

*Scoring*

For the WFT, the total number of correct animals listed in 1 minute was used as the final score for the WFT. For the CST, the average time to complete the motor condition was subtracted from the time in condition A, B and C. The final outcome measure was the time in seconds on condition C, minus the average of condition A + B (after correcting for motor speed). For the SCWT, the interference score was calculated by subtracting the time of card 2 from the time in seconds on card 3.

Raw scores for each subtest were then adjusted for sex, age, and education (classified according to the Dutch Verhage system (Verhage, 1964), which ranges from level 1 [less than six years of primary education] to level 7 [university degree]) by transforming them into Z-scores relative to population norms (Schmand et al., 2012; Van der Elst et al., 2006), yielding three EF Z-scores per patient. Since Z-score calculation may lead to uninterpretable outliers, especially when a patient is particularly slow, negative Z-scores were capped at Z = -2.5.

**Magnetoencephalography**

To aid visual inspection of the data, the cross-validation Signal Space Separation (SSS; (van Klink et al., 2017)) was applied, after which we (SDK, LD, LCB, MLMZ) removed at most 12 noisy channels. We used the temporal extension of SSS (Taulu & Simola, 2006) in MaxFilter (version 2.2.15) for further noise removal. Patients' head position was recorded continuously with head position coils, which were digitized together with the scalp shape using a 3D digitizer (Fastrak, Polhemus; Colchester, VT, USA). Anatomical MRI (GE Discovery 3T magnet, Milwaukee, USA; voxel size 1mm x 0.5mm x 0.5mm) was used for co-registration with the digitized scalp surface using a surface matching approach (estimated accuracy 4 mm (Whalen et al., 2008)). A single best-fitting sphere was fitted to the scalp outline and used as a volume-conductor model for the beamformer approach described below. We normalized the co-registered MRI to a template and, following inverse-normalization, labeled the voxels in the co-registered MRI according to the Automated Anatomical Labeling atlas (Tzourio-Mazoyer et al., 2002). Broadband time series of neuronal activity were then reconstructed for each region's centroid (Hillebrand et al., 2016) using a scalar beamformer approach (Hillebrand et al., 2012). The normalized (Cheyne et al., 2007) beamformer weights were based on the lead fields for dipolar sources, broadband (0.5-48Hz) data covariance, and unity noise covariance (for the estimation of the optimum orientation (Sekihara et al., 2004)).

Supplementary Figure 1. Individual changes in executive functioning at both time points

The raincloud plots with blue clouds, raindrops and lines represent the IDH-mutant non-codeleted and red the IDH-mutant 1p/19q-codeleted glioma subgroup. Scores below the dashed line at -1.5 indicate clinically relevant cognitive deficits.

Supplementary Figure 2. Individual changes in multilayer integration per glioma subtype

Each panel shows multilayer eigenvector centrality of the frontoparietal network in patients with IDH-mutant 1p/19q-codeleted, IDH-mutant non-codeleted, and IDH-wildtype gliomas at both time points.

Supplementary Table 1. Non-significant results concerning multilayer integration

| Dependent | adj. $R^2$ | F (df) | *p*-value | Corrected *p*-value | Included predictors | B (stand.) | *p*-value | Excluded variables | *p*-value |
|---|---|---|---|---|---|---|---|---|---|
| T1 Word fluency | 0.135 | 3.73 (2,33) | 0.035 | 0.104 | IDH-wildtype (ref = IDH-mutant, 1p/19q-codeleted) | -0.430 | 0.015 | T1 multilayer EC | 0.451 |
| | | | | | IDH-mutant, non-codeleted | -0.305 | 0.079 | Non-frontal tumor (ref = frontal) | 0.126 |
| T1 Inhibition | 0.060 | 3.25 (1,34) | 0.080 | - | IDH-wildtype | -0.295 | 0.080 | Non-frontal tumor | 0.709 |
| | | | | | | | | IDH-mutant, non-codeleted | 0.575 |
| | | | | | | | | T1 multilayer EC | 0.533 |
| Δ Inhibition | -0.013 | 0.652 (1,27) | 0.426 | - | RT (ref = no treatment) | -0.154 | 0.426 | Δ multilayer EC | 0.885 |
| | | | | | | | | RT and XT | 0.645 |
| Δ Word fluency | 0.396 | 9.84 (2,25) | <0.001 | 0.002* | IDH-wildtype | 0.482 | 0.005 | Δ multilayer EC | 0.827 |
| | | | | | IDH-mutant, non-codeleted | 0.626 | 0.001 | RT and XT | 0.519 |
| Δ Word fluency | 0.617 | 15.5 (3,24) | <0.001 | <0.001* | T1 Word fluency | -0.498 | 0.001 | T1 multilayer EC | 0.743 |

| | | | | | | | | | |
|---|---|---|---|---|---|---|---|---|---|
| | | | | | IDH-wildtype | 0.327 | 0.020 | RT | 0.946 |
| | | | | | IDH-mutant, non-codeleted | 0.491 | 0.001 | RT and XT | 0.195 |
| Δ Inhibition | 0.197 | 4.44 (2,26) | 0.022 | 0.066 | T1 Inhibition | -0.452 | 0.014 | T1 multilayer EC | 0.812 |
| | | | | | Age | -0.309 | 0.084 | RT and XT | 0.599 |
| | | | | | | | | RT | 0.167 |
| Δ Set shifting | 0.223 | 4.88 (2,25) | 0.016 | 0.049* | T1 Set shifting | -0.340 | 0.069 | T1 multilayer EC | 0.766 |
| | | | | | Active treatment at T2 (ref = no treatment) | 0.341 | 0.091 | Interval resection - NPA | 0.660 |

EC = eigenvector centrality, Δ = change score (T2-T1), RT = radiotherapy, XT = chemotherapy, NPA = neuropsychological assessment. In case of dummy-coded variables, the reference category (ref) is indicated at first mention only. * = significant p-value (<.05) after Bonferroni correction.

Supplementary Table 2. Regions of the frontoparietal network within the atlas

| Regions of the frontoparietal network |
|---|
| Inferior frontal gyrus, orbital part, L |
| Inferior frontal gyrus, orbital part, R |
| Superior frontal gyrus, L |
| Superior frontal gyrus, R |
| Middle frontal gyrus, L |
| Middle frontal gyrus, R |
| Superior parietal gyrus, L |
| Superior parietal gyrus, R |
| Inferior parietal gyrus, L |
| Inferior parietal gyrus, R |
| Middle cingulate gyrus, L |
| Middle cingulate gyrus, R |

Legend. L = left hemisphere, R = right hemisphere

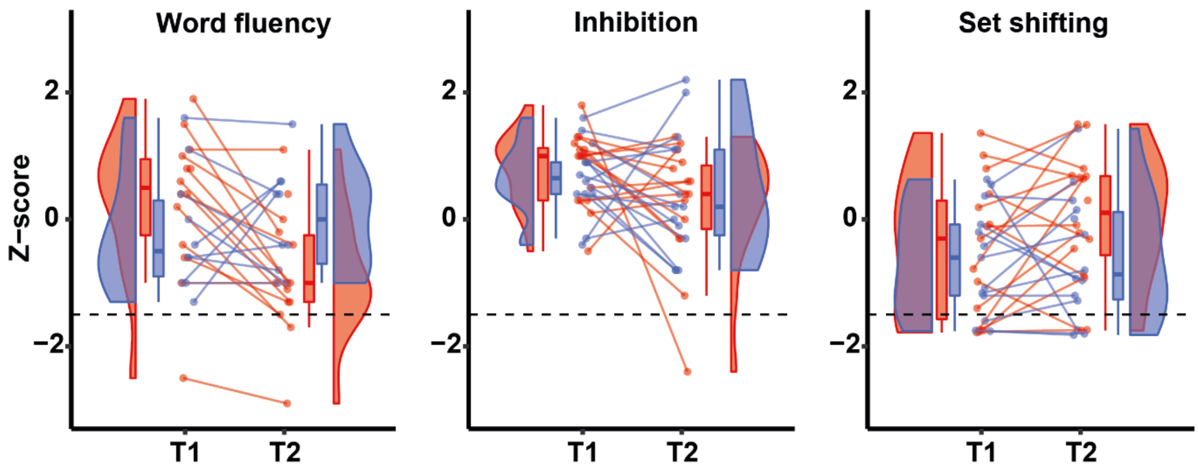


Supplementary Figure 1.

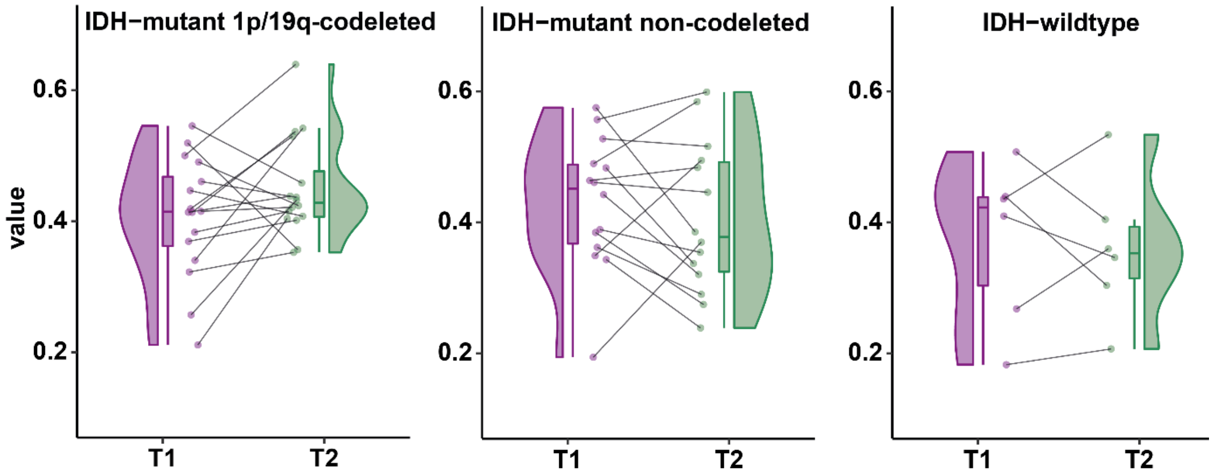


Supplementary Figure 2.
